# iCPET calculator: a web-based application to standardize the calculation of alpha distensibility in patients with pulmonary arterial hypertension

**DOI:** 10.1101/2023.02.21.23286277

**Authors:** Jordan Elliott, Nainika Menakuru, Kellan Juliet Martin, Farbod Nicholas Rahaghi, Franz P. Rischard, Rebecca R. Vanderpool

## Abstract

**Introduction:** Pulmonary vascular distensibility associates with right ventricular function and clinical outcomes in patients with unexplained dyspnea and pulmonary hypertension. Alpha distensibility coefficient is determined from a non-linear fit to multipoint pressure-flow plots. The study aims were 1) to create and test a user-friendly tool to standardize analysis of exercise hemodynamics including distensibility and 2) to investigate changes in distensibility following treatment in patients with pulmonary arterial hypertension.

**Methods:** Participants with repeat exercise right heart catherization and PAH were identified from the University of Arizona PH Registry (n=29). Single-beat analysis was used to quantify right ventricular function including the coupling ratio and diastolic stiffness. Prototypes of the *iCPET calculator* were developed using Matlab, Python and RShiny to analyze exercise hemodynamics and alpha distensibility coefficient, α (%/mmHg) from multi-point pressure flow plots. Interclass coefficients were calculated for inter-platform and interobserver variability in alpha.

**Results:** No significant bias in the intra-platform (Matlab vs RShiny: ICC: 0.996) or inter-observer (ICC: 0.982) comparison of alpha values. Participants with PAH had a significant decrease in afterload at follow-up (p<0.05) but no significant change in alpha distensibility. At follow-up, participants with a resting mean PA pressure < 25 mmHg had no change in pressure, resistance or alpha distensibility. Alpha distensibility significantly correlated with PA compliance at both the index and follow-up visit.

**Discussion:** The *iCPET calculator* standardizes alpha distensibility calculations. In this retrospective cohort, alpha distensibility did not change despite a decrease in pulmonary vascular afterload (PVR and mPAP) at follow-up after treatment with pulmonary vasodilators.

## INTRODUCTION

The right ventricle (RV) has considerable reserve in the face of increased afterload but decreased RV function strongly associates with mortality in patients with pulmonary arterial hypertension (PAH).(1) RV function measurements including ejection fraction at rest must decrease more than 40% before there is a rapid progressive dilation in the RV, poor prognosis and RV failure symptomatology. Invasive cardiopulmonary exercise testing (iCPET) is an important method for quantifying RV function under stressed conditions that is not possible with current resting measurements (2–6), enabling detection of right ventricular dysfunction earlier in the disease course. The RV contractile reserve (RVRC) are objective measurements of the ability of the RV to adapt to altered afterload and stresses that can be quantified by a change in PA pressure, heart rate, cardiac output, RV contractility and RV-PA coupling ratio (7–11).

Exercise hemodynamics also allow for the opportunity to also assess the pulmonary circulation the pulmonary vascular distensibility coefficient (α) using the Linehan distensible model of the pulmonary circulation. (12–15) Studies into pulmonary vascular distensibility (α) have focused on defining the upper limits of normal (14,15), early detection of pulmonary vascular disease(15–17), or the differential diagnosis of PAH from left sided heart failure(18). In a comprehensive paper, Malhotra et al.(18) demonstrated decreased pulmonary vascular distensibility in patients with PAH (0.4%) compared to patients with normal pulmonary pressure (1-2%) or heart failure (0.8-0.9%). Pulmonary vascular distensibility is modifiable by vasodilatory therapy in exercise induced PH(17) and heart failure patients. (18) Pulmonary vascular (PV) distensibility has been found to associate with outcomes in patients with borderline PH and heart failure ejection fraction. (18,19). In a comprehensive study in patients with PAH, exercise-induced PH and heart failure preserved ejection fraction, PV distensibility was an independent predictor of peak RV-PA coupling. (11) RV diastolic stiffness, another measure of RV function, significantly associates with outcomes in patients with pulmonary hypertension (20) and is sensitive to three months of prostacyclin therapy in patients with PAH. (2)

While the clinical relevance of the pulmonary vascular distensibility coefficient has been demonstrated, the implementation requires clinical practitioners to use a complex set of markers in the analysis of exercise hemodynamics. An additional barrier to implementation is that the Linehan model of pulmonary vascular distensibility is a non-linear equation (Eq. 1) that requires custom programing in Excel, Matlab, R or another data analysis program. Solving for the alpha distensibility coefficient α has led to varying implementations of the equation and analysis across centers (17,21,22). Recent efforts have been made to standardize invasive cardiopulmonary exercise testing across centers in the Pulmonary Vascular Disease Phenomics (PVDOMICS) study. (23) The next step is to standardize the calculation of the pulmonary vascular distensibility coefficient and ultimately improve the quality and reproducibility of this analysis. Commercial and non-commercial software packages for the analysis of the distensibility coefficient are not available. In this paper, we introduce the *iCPET Calculator* that is a multi-platform web-based exercise hemodynamics software tool. The calculator is designed to be user friendly with a graphical user interface (GUI) that allows the user to input, modify, visualize, and save exercise hemodynamics using open-source tools. Considerations in the development of the calculator included the cost of the development platform (MATLAB, Python, and R), accessibility of the tool to non-programmers, and the speed of the analysis. (24,25)

Using this web-based analysis tool, we tested the hypothesize that decreased pulmonary vascular distensibility associates with reduced right ventricular function and predicts therapeutic response in patients with pulmonary arterial hypertension. To do this, we investigated the therapeutic effect of PAH medication on pulmonary vascular distensibility in patients with PAH who had serial assessment of exercise hemodynamics. Specifically, we investigated associations between the change in pulmonary vascular distensibility with the change in a) systolic RV function, b) diastolic RV function.

## METHODS

### Study Population

Participants were identified from the University of Arizona Pulmonary Hypertension registry (UA PH registry) that had a right heart catheterization and an invasive cardiopulmonary exercise test (iCPET, n = 30 participants, RHC date of procedures between 2011-2018).

Additionally, the participant needed at least two iCPETs with at least two stages of exercise in addition to the resting RHC measurements (**Figure 1A**). Completeness of the RHC and iCPET data was evaluated and participants were excluded if they were missing key exercise data needed for the alpha distensibility calculations like exercise wedge pressure, cardiac output or mean PA pressure (n = 1 participant). This is a cross-sectional analysis, and the earlier visit was labeled as the **Index** time point and the subsequent visit was labeled as the **Follow-up** time point. The pulmonary hypertension diagnosis for the participants was verified by a physician based on the world symposium of pulmonary hypertension (WSPH) guidelines.(26) Participants had a mix of pulmonary hypertension classification and the current analysis focused on those with a mean PA pressure less than 25 mmHg (mPAP < 25 mmHg) and those with WSPH group 1 pulmonary hypertension (PAH) based on 2015 ESC/ERS PH Guidelines. (27) One participant with WSPH group 4 PH was excluded from the analysis (**Figure 1A**). All participants gave informed consent through the Respiratory-Related Disease Patient Registry (UA PH registry), which was approved by the institutional review board at the University of Arizona (IRB Protocol no. 1100000621).

**Figure 1.**
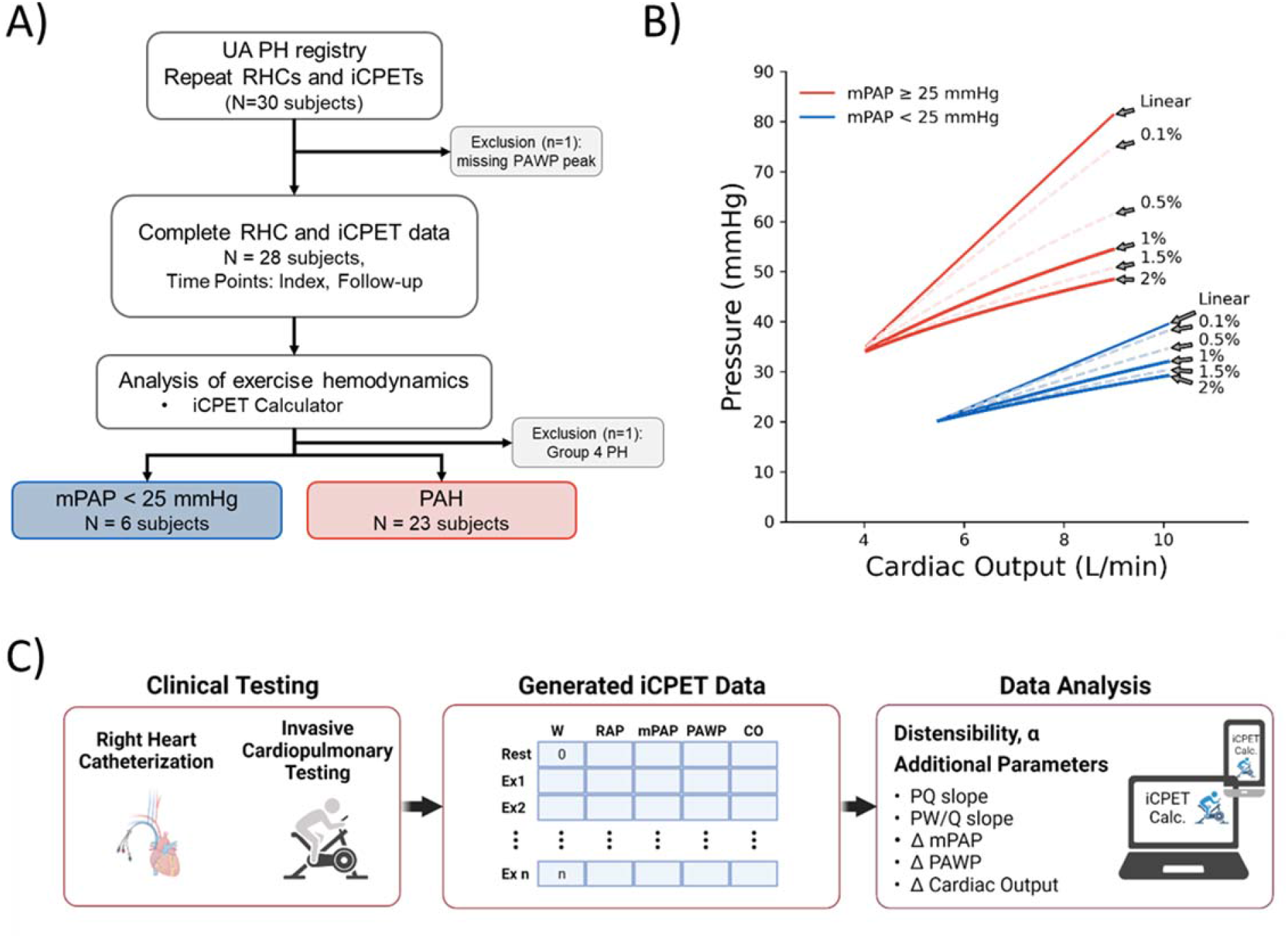
Analysis of exercise hemodynamics and alpha distensibility in patients with and without pulmonary hypertension (PH). **A)** Identification of participants from the university of Arizona PH registry that had repeat right heart catheterizations (RHC) that included invasive cardiopulmonary exercise testing (iCPET) at both the index and follow-up visit. **B)** Theoretical pressure-flow relationships at different alpha distensibility in patients with a mean PA pressure of 25 mmHg or more (mPAP ≥ 25 mmHg) and less than (mPAP < 25 mmHg). **C)**The generated iCPET hemodynamic data were analyzed for alpha distensibility as well as other metrics including the slope of the pressure-flow relationship for PA pressure (PQ slope) and wedge pressure (PW/Q slope) and the exercise-induced change in mean PA pressure (ΔmPAP), wedge pressure (ΔPAWP) and cardiac output (ΔCO).

### Invasive Cardiopulmonary Exercise Testing

Right heart catheterizations (RHC) were performed in accordance with clinical guidelines. Briefly, a pulmonary artery catheter was inserted into the antecubital vein and advanced into the right atrium, right ventricle, and into the pulmonary artery. Resting pressure including right atrial pressure (RAP), right ventricular pressure (RVP), mean pulmonary arterial pressure (mPAP) and pulmonary wedge pressure (PAWP). Cardiac output (CO) was measured by thermodilution and direct Fick method.

Following the resting RHC, participants underwent a symptom-limited incremental CPET with invasive pulmonary pressure and cardiac output. Work rate was increased every 2 min following a step protocol with steps of 5, 10, 15, or 20W depending on the patient’s functional status. Peak exercise was determined either by a peak respiratory exchange ratio of > 1.10 or symptom limitations. Breath-by=breath pulmonary gas exchange was recorded continuously to determine O_2_ consumption during exercise. In the last minute of each exercise stage, mPAP and PAWP were recorded (Xper Cardio Physiomonitoring System; Phillips) and blood samples were drawn from the PA catheter to measure hemoglobin and O_2_ saturation. Cardiac output was calculated by the direct Fick method (cardiac output=O_2_ consumption/arteriovenous O_2_ difference) where systemic O_2_ saturation from pulse oximetry was used in the arteriovenous O_2_ difference (arteriovenous O_2_ difference=systemic–PA O_2_ content).

### Pulmonary Vascular Distensibility and Compliance

Pulmonary vascular distensibility, a measure of distal vascular distensibility and pulmonary vascular compliance, a measure of global compliance, were both determined to quantify pulmonary vascular mechanics. Hemodynamics from the multi-stage invasive cardiopulmonary exercise testing were used to estimate the pulmonary vascular distensibility coefficient α using the Linehan distensibility model

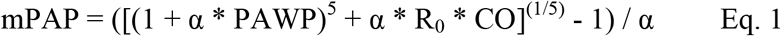

where mean PA pressure (mPAP) is a function of distensibility coefficient (α), wedge pressure (PAWP), cardiac output (CO) and unstressed resistance (R_0_). (22,28) R_0_ is estimated by the ratio of mPAP and cardiac output at rest (mPAP/CO). Theoretical pressure-flow relationships demonstrate the effect of alpha distensibility on the developed pressure at higher cardiac outputs (**Figure 1B**).

Pulmonary arterial (PA) compliance was calculated as the ratio of stroke volume to pulmonary artery pulse pressure (PACa = SV/(systolic PAP–diastolic PAP). Stroke volume (SV) was determined as the ratio of cardiac output and heart rate (SV=cardiac output / heart rate). Pulmonary vascular resistance (PVR) was calculated as the ratio of (mPAP – PAWP) and cardiac output. The pulmonary vascular resistance-compliance relationships were investigated and the RC-time constant was calculated as the product of PACa and PVR.

### Right Ventricular Function

Single-beat analysis was used on RV pressure waveforms to assess right ventricular function. (29–31). Max isovolumetric pressure (Pmax) was determined by fitting a sinusoidal curve to the early isovolumetric contraction and late isovolumetric relaxation regions of the RV pressure waveform. An algorithm was used to determine these regions defined by the first and second derivate of the pressure with respect to time (29). Only minor manual modifications of these regions were allowed to optimize the sinusoidal fit. End-systolic pressure was estimated from mean PA pressure using the proposed equation ESP = 1.65*mPAP – 7.76 based on invasive conductance catheter measurements. (31) The end systolic elastance (Ees) was calculated as (Pmax-ESP)/Stroke Volume. Arterial elastance (Ea) was calculated as the ratio of ESP to stroke volume leaving the coupling ratio (Ees/Ea) as the ratio of (Pmax-ESP)/ESP. RV stroke work index (RVSWI) was calculated as 0.0136 ∙ SVI ∙ (mPAP – RAP) in units of g/m/beat/m^2^.

### iCPET Calculator Program Design

Invasive cardiopulmonary exercise tests generate a lot of data to be analyzed including workload, right atrial pressure, pulmonary artery pressure, wedge pressure and cardiac output. Additional clinically meaningful parameters including the pressure-flow slopes and incremental change in pressure require additional calculations (**Figure 1C**). The development of the iCPET calculator by the team of engineers, pulmonologists, and physiologists focused on improving the analysis and interpretation workflow for iCPETs. Program design considerations included 1) the need for a graphical user interface to enter, review and modify data; 2) the capability to generate the α distensibility coefficient and other hemodynamic parameters with one click; 3) the ability to save analysis and figures; 4) keeping the analysis local to protect patient data; 5) the ability to make the program platform agnostic and run without the requirement of expensive licenses or large support files; and 6) provides added value to current practices (**Table S1 and Figure S1**). Modern data analysis programs have the tools and packages necessary to create such a program. The *iCPET calculator* was prototyped in four analysis solutions including Excel, Matlab, Python and R/Rshiny. The Matlab and RShiny versions of the *iCPET calculat*or were used for repeated analysis of pulmonary hemodynamics in the identified patient cohort from the UA PH registry. Bland-Altman and Intraclass Correlation Coefficient (ICC) analyses were used to compare the calculation of distensibility, α, between the Matlab and RShiny versions of the *iCPET calculator* to determine the effect of different implementations of the non-linear fit to the measured data.

### Statistical Analysis

Results are expressed as mean ± standard deviation (SD) or median [interquartile range (IQR)]. A p-value < 0.05 was considered significant. Comparisons between groups were made using a t-test or Mann–Whitney test, where appropriate. Repeated-measures analysis was performed to asses changes over time (index, follow-up) between the two groups (mPAP < 25 and PAH). The lmer function in the lme4 package in R was used to construct repeated-measures linear mixed-effect models. Correlations between variables were calculated using Pearson’s correlation coefficient or Spearman rank correlation coefficient, where appropriate. The statistical analyses were performed using R programming language (r-project.org, version 4.1.1) and RStudio (version 1.4.1717).

## RESULTS

Twenty-eight participants were identified with complete RHC and iCPET data at an index and follow-up time point with a median follow-up time of 19 [8-28] months. Participants with a mean PA pressure less than 25 mmHg (mPAP < 25) had an average age of 67 ± 5 years and a median follow-up time of 30.1 [16.2-42.6] months (**Table 1**). They were female (80%), non-Hispanic with an average BMI of 34 ± 16 kg/m^2^. Participants with PAH had an average age of 54 ± 13 years and a median follow-up time of 16.1 [5.8-23.8] months. They were predominately female (87%) and Non-Hispanic (86%).

**Table 1.** Demographics and Baseline Characteristics in patients with pulmonary arterial hypertension (PAH) and without pulmonary hypertension (Controls) at their index and follow-up right heart catheterization visit.

|  | mPAP < 25 (n = 6) |  | PAH (n = 22) |  |
| --- | --- | --- | --- | --- |
| Visit | Index | Follow-up | Index | Follow-up |
| Female (%) | 5 (80) |  | 19 (87) |  |
| Hispanic (%) | 0 (0) |  | 3 (14) |  |
| Deaths (%) | 0 (0) |  | 2 (9) |  |
| Age (years) | 67 ± 5 |  | 54 ± 13 |  |
| Time between visits (months) |  | 30.1 [16.2 – 42.6] |  | 16.1 [5.77 – 23.8] |
| BMI (m<sup>2</sup>) | 34 ± 16 | 28 ± 6 | 29 ± 5 | 28 ± 5 |
| RA pressure (mmHg) | 4 ± 3 | 3.5 ± 1 | 4 ± 3 | 3.5 ± 3 |
| RV ESP (mmHg) | 23 ± 6 | 22 ± 4<sup>a</sup> | 70 ± 13<sup>a,b</sup> | 54 ± 14<sup>a,b,c</sup> |
| Ees (mmHg/ml) | 0.60 ± 0.43 | 0.62 ± 0.25 | 1.41 ± 0.76<sup>a,b</sup> | 1.15 ± 0.58<sup>b</sup> |
| Ea (mmHg/ml) | 0.33 ± 0.12 | 0.29 ± 0.10 | 1.15 ± 0.45<sup>a,b</sup> | 0.88 ± 0.43<sup>a,b</sup> |
| Ees/Ea | 1.89 ± 1.22 | 2.54 ± 1.51 | 1.25 ± 0.55<sup>a,b</sup> | 1.46 ± 0.82<sup>b</sup> |
| Eed (mmHg/ml) | 0.25 ± 0.18 | 0.26 ± 0.15 | 0.80 ± 0.51<sup>a,b</sup> | 0.55 ± 0.21<sup>b</sup> |
| RVS WI (g/m/beat/m<sup>2</sup>) | 8.0 ± 2.4 | 8.8 ± 2.4<sup>a</sup> | 20.6 ± 5.8<sup>a,b</sup> | 16.1 ± 5.1<sup>a,b,c</sup> |

BMI: Body Mass Index, RA: Right atrium, RV ESP: Right ventricular end-systolic pressure, Ees: end-systolic elastance, Ea: Arterial elastance, Ees/Ea: Coupling Ratio, Eed: End-diastolic elastance. RVS WI: RV stroke work index. Data presented as mean ± standard deviation, median [interquartile range] or number (percent). <sup>a</sup> vs mPAP<25 Index, <sup>b</sup> vs mPAP<25 Follow-up, <sup>c</sup> vs PAH index, and <sup>d</sup> vs PAH Follow-up.

### iCPET Calculator to analyze Exercise Hemodynamics

In this this project, we focused on the four very commonly available platforms of Excel, Matlab, Python and R/Rshiny for the development of an *iCPET calculator*. A more detailed discussion of the design considerations is included in **Table S1** and details of the development in each platform are included in **Figure S1**. The current version of the *iCPET calculator* was designed to be usable on a desktop or laptop computer and on mobile devices using the R programming language and the RShiny Package (an R development tool for the development of the graphical user interface) (**Figure 2**). The user interface has a linear end-user experience that allows the user to start with the participant information and then enter the invasive hemodynamics into the RHC/iCPET stages table. After selection of the rest and exercise stages, the program analyzes alpha distensibility and determines the slope of the pulmonary artery, wedge and right atrial (RA) pressure-flow relationships. The RA pressure-flow relationship calculations are optional.

**Figure 2.**
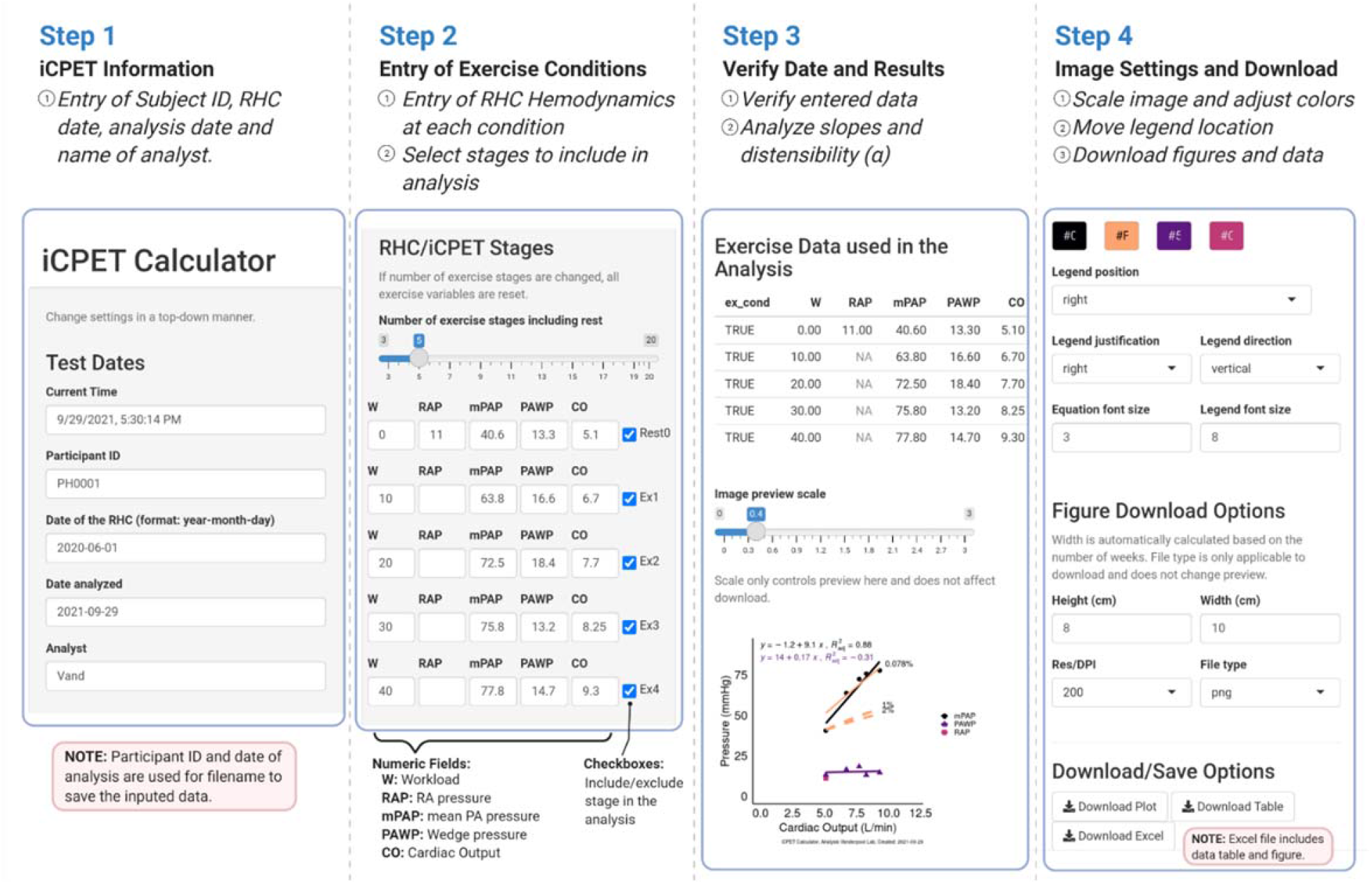
Screenshots of the iCPET calculator on a mobile device. The iCPET calculator allows the user to enter the measured exercise hemodynamics and then calculate the slopes of the pressure-cardiac output relationships and alpha distensibility. The steps to use the app include: S**tep 1:** The user enters information about the test including an identifying tag, date of the RHC/iCPET, date of the analysis and the name of the analyst. **Step 2:** The number of exercise stages including rest are selected and then the workload (W) and pressure data are entered into the resulting table. To increase flexibility, check boxes on the right-hand side are used to select exercise stages to be included in the calculations. **Step 3:** The entered data is displayed as a table and a figure to verify the entered results are correct. The figure allows the user to visually assess the pressure-flow relationships for mean PA pressure (mPAP), wedge pressure (PW) and RA pressure (RAP). Entering the RA pressure is optional. **Step 4:** The user can adjust settings for the figure including colors and legend location before selecting how to download and save the analysis. The downloaded Excel file will include the test information (Patient ID, date of RHC, date analyzed and analyst), exercise data from the entered table, alpha distensibility, and slopes of the pressure-flow relationships including the PQ slope and PW/Q slope.

Key components of the program are 1) the user’s ability to verify and edit entered data in an interactive fashion and 2) the ability to save the analysis tables, figures and resulting analysis for later review and analysis. To assess the stability of the alpha distensibility calculations, the results from the Matlab and Rshiny analysis programs were compared to determine inter-platform variability (**Figure 3A**). There were no biases in the calculated alpha distensibility (Bias: -0.002 %/mmHg, LOA: -0.06 to 0.05 %/mmHg). The inter-observer variability in the calculation of alpha distensibility using the RShiny app showed no biases in the calculated alpha distensibility (Bias: -0.008 %/mmHg, LOA: -0.115 to 0.099 %/mmHg) (**Figure 3B**).

**Figure 3.**
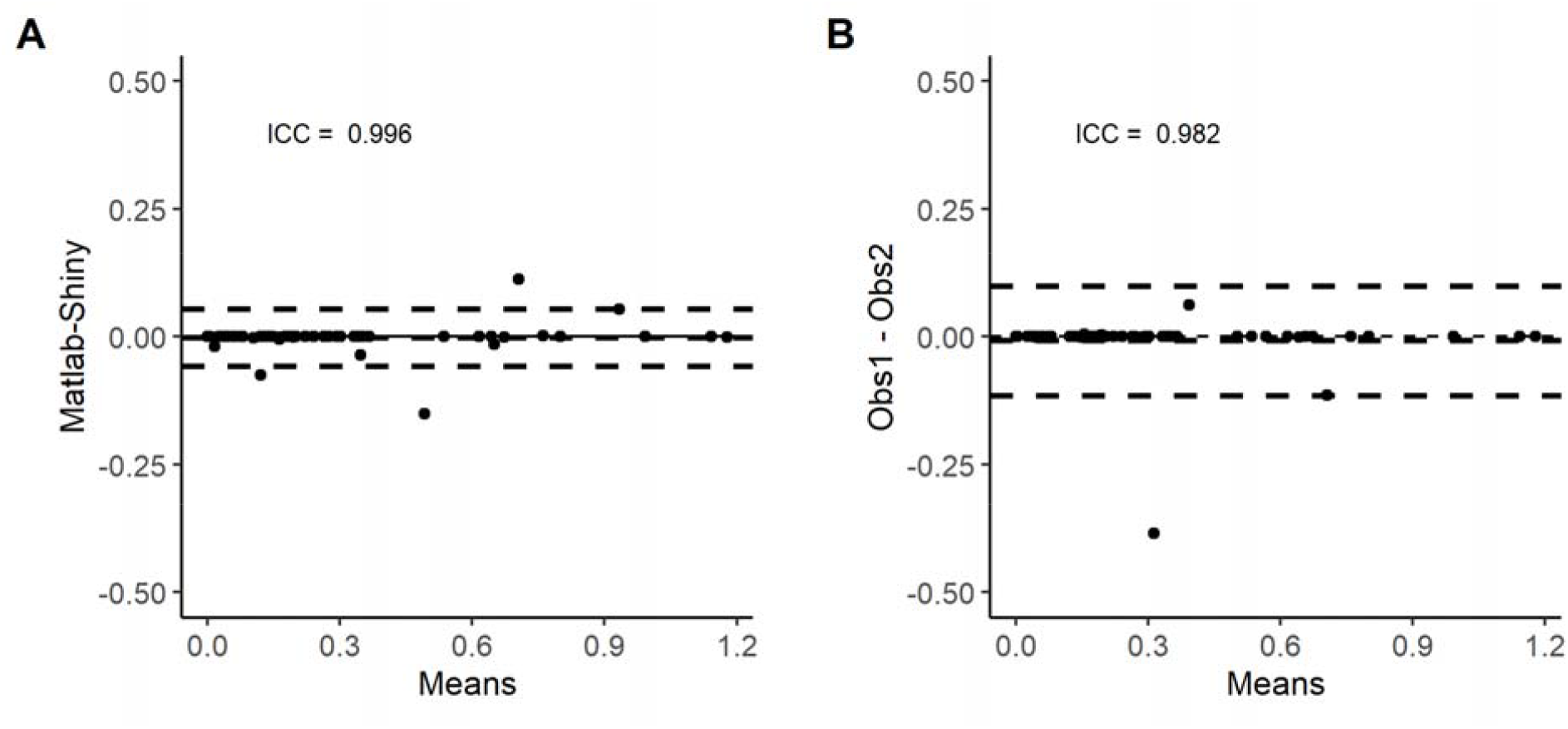
A) Inter-platform variability and B) inter-reader variability in the calculation of alpha distensibility.

### Changes in Resting Right Ventricular Function

Pulmonary vascular afterload decreased in participants with PAH between index and follow-up visits as measured by decreased mean PA pressure (**Figure 4A**) and pulmonary vascular resistance (**Figure 4B**). There was a significant decrease in arterial elastance, a non-significant decrease in Ees and resulting in no change in the coupling ratio Ees/Ea in the PAH group (**Table 1**). There were no significant changes in participants with a mean PA pressure less than 25 mmHg. RV stroke work index decreased in the participants with PAH but there were no significant changes in RV diastolic stiffness at follow-up in both groups (**Table 1**).

**Figure 4.**
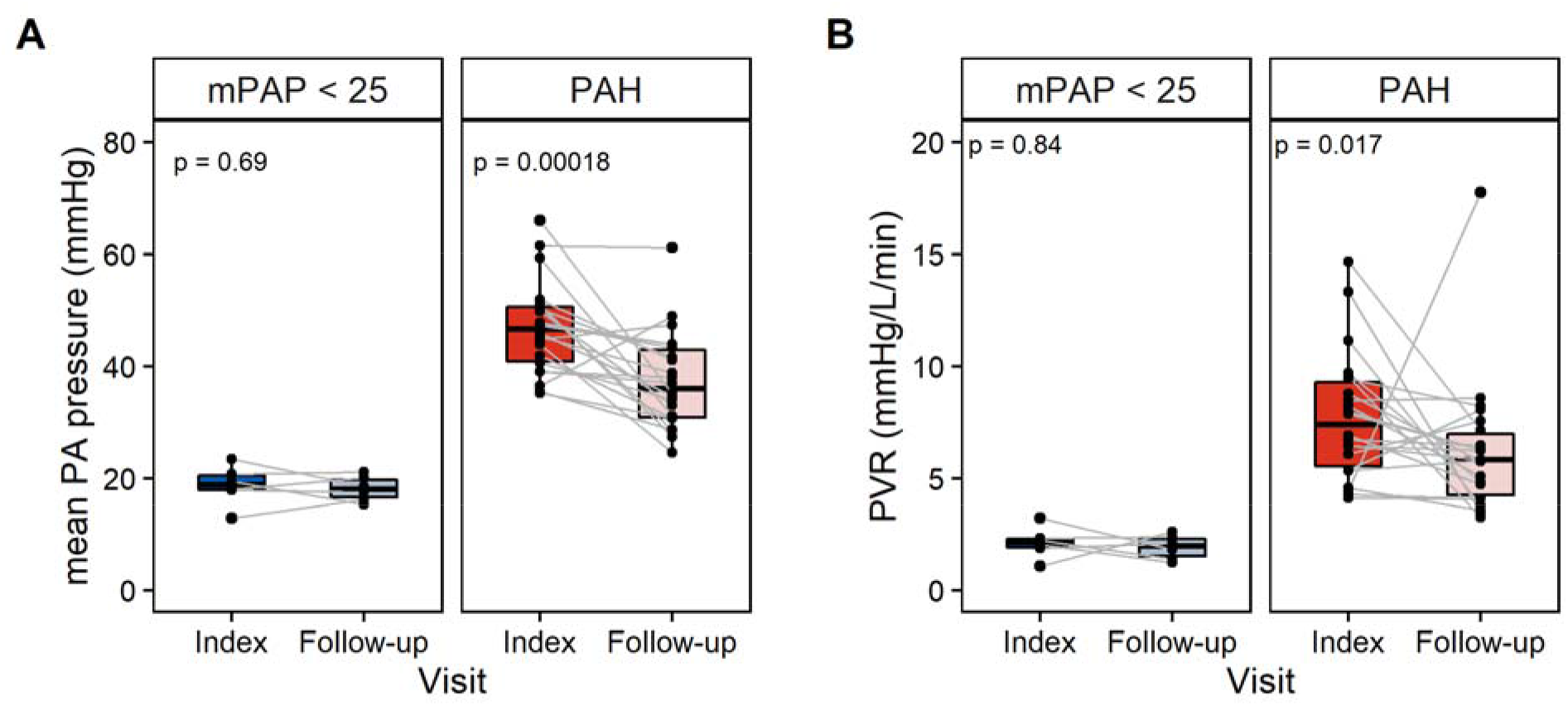
Decreased afterload in patients with pulmonary arterial hypertension (PAH) at follow-up. **A)** Patients with PAH have decreased mean PA pressure at follow-up. There were no significant changes in patients without pulmonary hypertension (mPAP < 25 mmHg). **B)** Decreased pulmonary vascular resistance (PVR) in patients with PAH. There were no significant changes in patients with mPAP < 25 mmHg.

### Exercise hemodynamics

Similar workloads were achieved in patients with mPAP < 25 mmHg or PAH at their index or follow-up RHC (**Table 2**). In the patients with PAH at their index visit, they achieved a median workload of 40 [30-44] W with an exercise-induced increase in heart rate of 34 ± 13 bpm, systolic PA pressure of 31 ± 15 mmHg, cardiac index of 1.8 ± 1.1 L/min/m^2^ resulting in a change of stroke volume index of 5 ± 9 ml/m^2^. There was no significant exercise-induced change in PVR or PA compliance. The slope of the mean PA pressure-flow relationship (mPAP/Q slope) was 5.9 [4.1-8.6] mmHg/L/min. At the follow-up visit, patients with PAH had similar exercise-induced increase in sPAP, cardiac output and stroke volume. There was no significant change in the mPAP/Q slope at follow-up. In the participants with mPAP < 25 mmHg, the peak workload was 37 [35-51] W with an exercise-induced increase in HR of 30 ± 13 bpm, sPAP of 17 ± 9 mmHg, cardiac index of 2.4 ± 1.4 L/min/m^2^ and stroke index of 11 ± 8 ml/m^2^. At follow-up, there were no significant changes compared to the index visit. In comparison to participants with mPAP < 25 mmHg, participants with PAH had increased PVR, mPAP/Q slope and decreased PA compliance (**Table 2**). Participants with PAH had more of an exercise induced increase in sPAP (ΔsPAP) compared to the mPAP < 25 mmHg group.

**Table 2.**
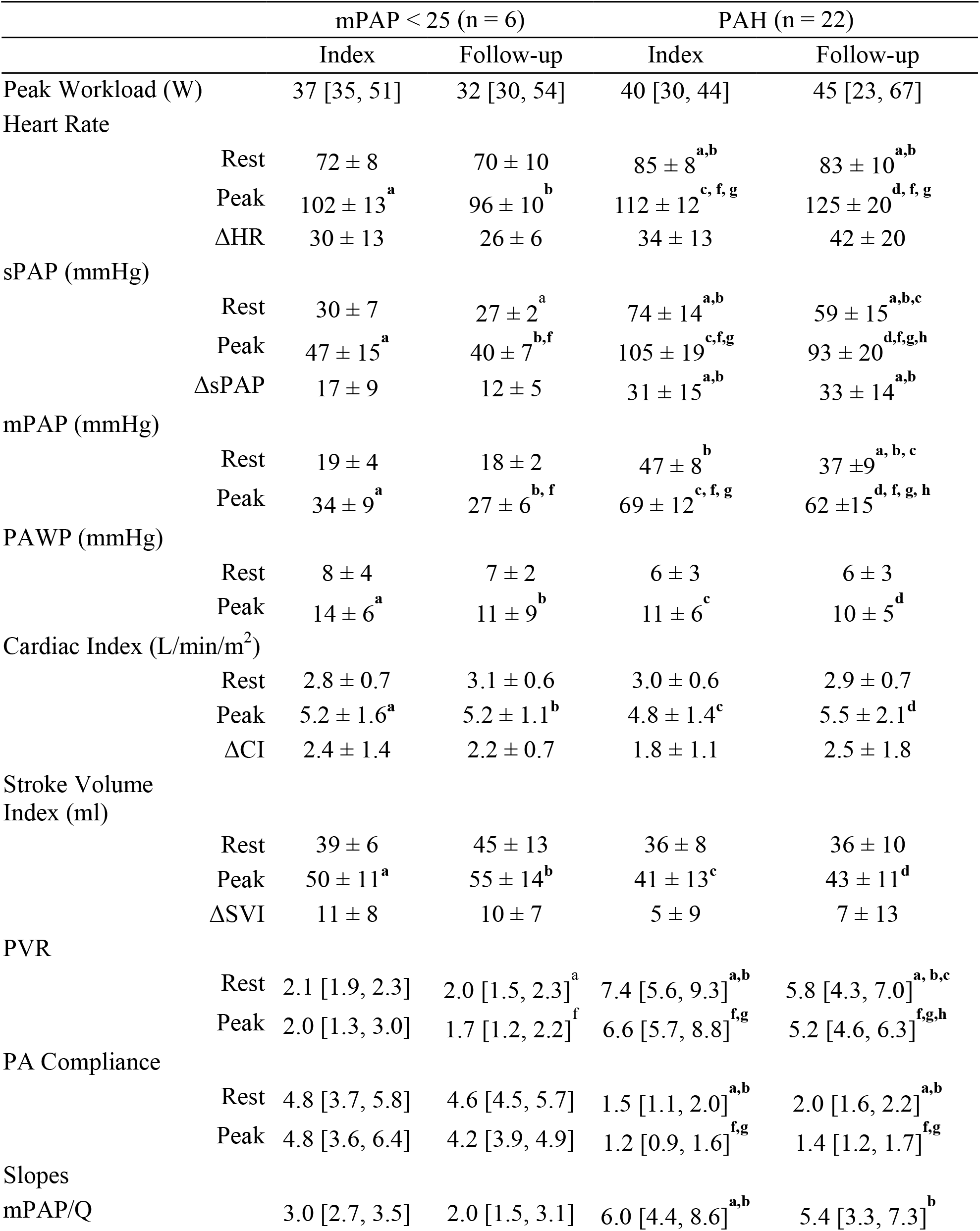
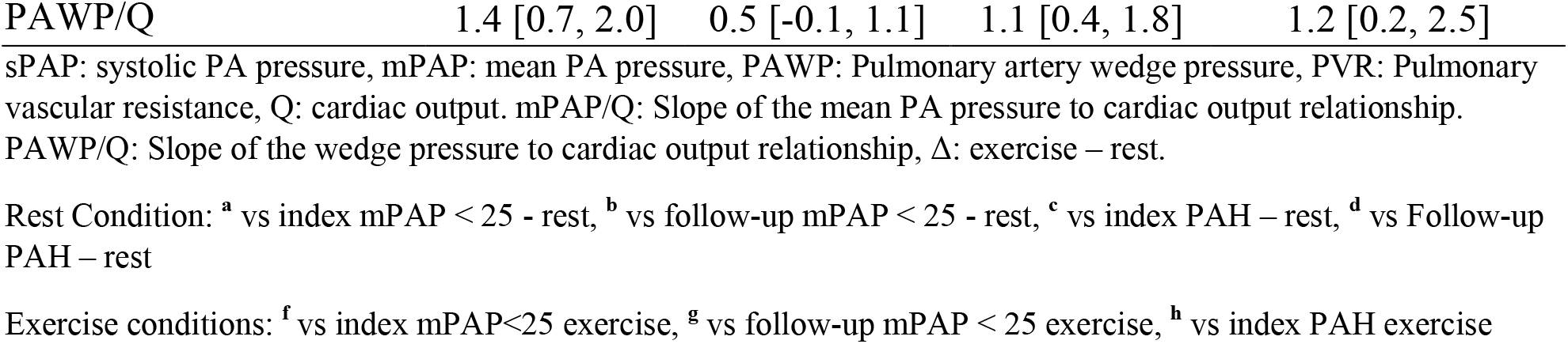
Exercise hemodynamics at index and follow-up invasive cardiopulmonary exercise test.

### PA Compliance and Alpha Distensibility

At the index RHC, exercise had minimal effect on the PA-compliance-PVR relationship (**Figure 5A)**. There was a small shift down and to the left of the exercise resistance-compliance curve (Figure 5A, dashed line, Ca__EX_ = 9.12/(0.22 +PVR__EX_)) compared to rest (Figure 5A, solid line, Ca__REST_ = 12.16/(0.6+PVR__REST_) at the index time point. From **Table 2**, there were no significant exercise-induced changes in PA compliance or PVR at the index visit. At follow-up, PVR significantly decreased in participants with PAH (**Table 2**) but PA compliance did not significantly change due to the resistance-compliance relationship and the high index PVR. There were no significant changes in the rest resistance-compliance relationship (**Figure 5B**) and the exercise-induced changes in PA compliance or PVR. RC time constant did not change between the index and follow-up visiting in both groups (**Figure 5C**). At baseline, Alpha distensibility was significantly decreased in the participants with PAH compared to participants with an mPAP < 25 mmHg (0.13 [0.04-0.25] %/mmHg vs 0.70[0.39-0.76] %/mmHg, respectively, p < 0.001). There was no significant change in alpha distensibility at follow-up in both groups (**Figure 5D**). Alpha distensibility significantly correlated with PA compliance at the index (**Figure 5E**) and follow-up (**Figure 5F**) right heart catheterization.

**Figure 5.**
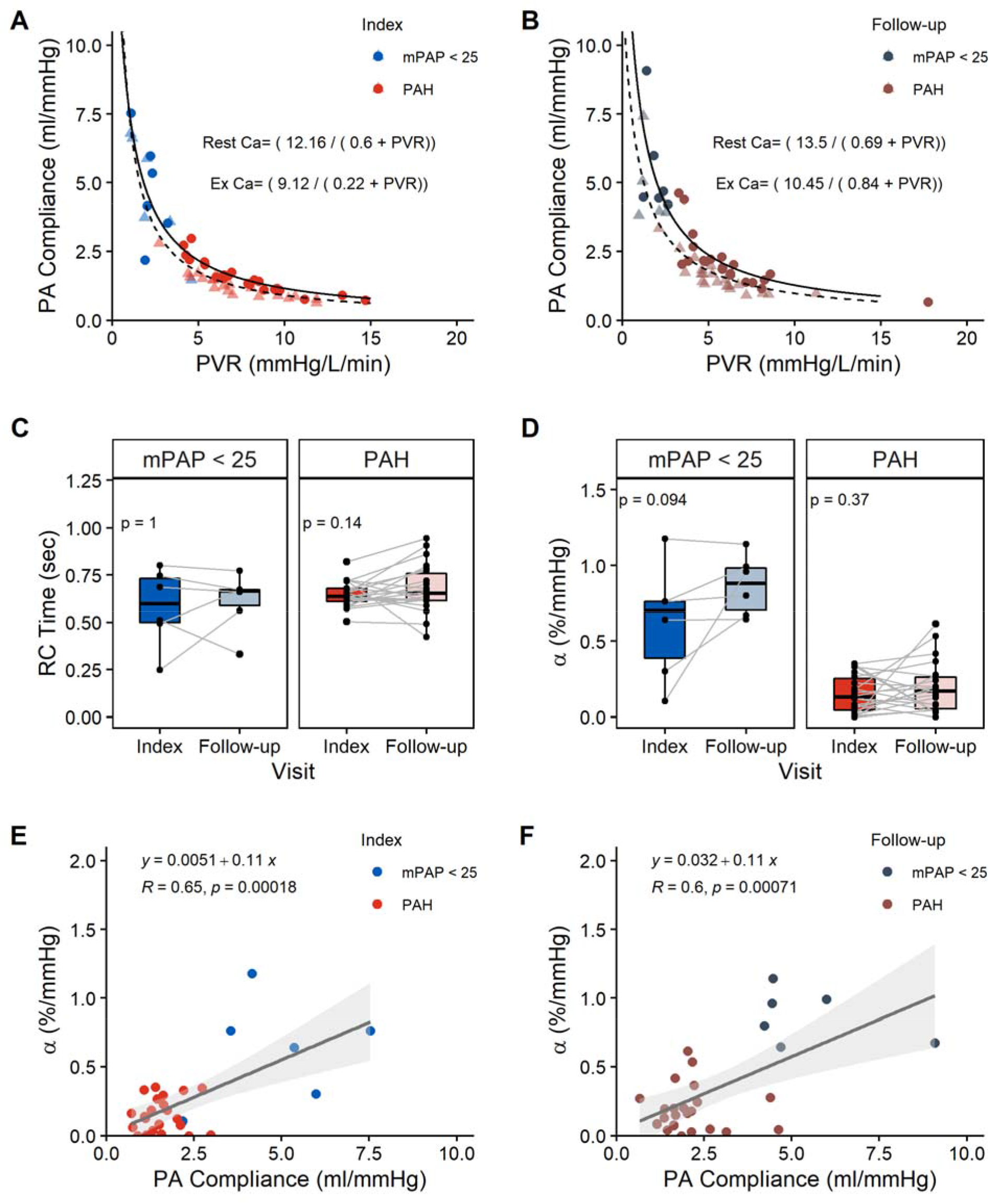
Alpha distensibility in context to the resistance-compliance relationship, PA compliance and RC-time. **A)** Inverse relationship between PA compliance and pulmonary vascular resistance (PVR) in patients with PAH (PAH) and without PH (Control) at rest (circle) and peak exercise (triangle) at the index time point. The exercise resistance-compliance curve (dashed line) had a small shift down and to the left compared to rest (solid line). **B)** Resistance-Compliance relationship at the follow-up time point. The exercise resistance-compliance curve (dashed line) had a shift down and to the left compared to rest (solid line). **C)** No significant differences in RC-time between index and follow-up RHC in both Control and PAH groups. **D)** No significant change in alpha distensibility (α) between index and follow-up visit for patients with PAH and without PH. PA compliance significantly correlates with alpha distensibility at the index **(E)** and follow-up **(F)** time point.

## DISCUSSION

In this study, we developed a web-based analysis tool, *iCPET Calculator*, to analyze invasive cardiopulmonary exercise tests and standardize the application of the distensible vessel model (28) in patients with pulmonary hypertension. A retrospective cohort of participants was identified and follow-up changes in alpha distensibility were investigated. The tool was successfully deployed to calculate key hemodynamic parameters including distensibility with good agreement between the different users and implementations. No differences in alpha distensibility were noted between index and follow up. Several factors could have contributed to this finding, including age of participants and the severity of PAH in the cohort.

### Standardizing analysis of alpha distensibility with the iCPET calculator

Calculation of alpha distensibility and the incremental slopes of the mPAP/Q and PAWP/Q slopes provide clinically meaningful information in the analysis and interpretation of invasive cardiopulmonary exercise testing. Four platforms (Microsoft Excel, Matlab, Python and RShiny) were considered in the development of this analysis tool (**Table S1**). Despite wide availability and user familiarity, a reason why Excel was excluded as an option was because the user would need to have optimization specific vocabulary to install and use the ‘Solver’ add-on. Determination of alpha in Excel also requires the user to have a relatively accurate guess for alpha distensibility to get an accurate result from the Solver add-on. Reproducibility of the analysis steps are difficult in Excel because the user analysis steps are not saved for further review. The other three platforms (Matlab, Python and Rshiny) all have powerful optimization packages and the ability for the development of application specific graphical user interfaces that are specific to analyze exercise hemodynamics. These custom programs require minimal user input. Analysis tools developed in Matlab and Python still required the user to download and install additional programs and environments. The web browser-based application for the *iCPET calculator* in R as an RShiny application allows users to use the calculator without needing to install additional software. RShiny applications have been beneficial in the analysis of RNAseq data with the development of a browser-based application for differential expression analysis (24). The flexibility of RShiny framework as a web-based program provides cross-platform compatibilities along with the excellent data visualization. It is possible to add in additional features and analysis in the future as new clinically important exercise hemodynamic variables are identified. The *iCPET calculator* provides a workflow that helps to reduce user errors and standardizes alpha distensibility calculations.

### Changes over time in mean PA pressure/flow slopes

Increased age and severity of pulmonary vascular remodeling have both been found to significantly impact the mean PA pressure/cardiac output (mPAP/Q) slope. Definitions of exercise pulmonary hypertension were re-introduced in the 2022 ESC/ERS PH guidelines as a mPAP/Q slope of > 3 mmHg/L/min. (32) Individuals with elevated mPAP/Q slopes have increased risk for future cardiovascular or death events.(34) Age is another factor that impacts the upper limit of normal for the mPAP/Q where the upper limit of normal decreases to 2.73 mmHg/L/min in individuals over the age of 50 years. (33) In our study, the participants with a mPAP <25 mmHg do not have a “normal” pulmonary vasculature with a mPAP/Q slope of 3.0 [IQR: 2.7-3.5] mmHg/L/min. At follow-up, there was no significant change in the mPAP/Q slope (2.0 mmHg/L/min) where the older age of the participants in this group (67 ± 5 years) may have contributed to these results. For the participants with PAH, they had elevated mPAP/Q slopes (6.0 [4.4-8.6] mmHg/L/min) that are consistent with significant pulmonary vascular remodeling. At follow-up in the PAH group, there was not a significant change in mPAP/Q slopes.

### Changes over time in pulmonary vascular stiffness and distensibility

Recent papers have demonstrated the functional relevance of alpha distensibility in patients with mild or exercise induced pulmonary hypertension. In a normal range of alpha distensibility (1-2%), the vasculature is able to expand to accommodate the increase in cardiac output with minimal increase in pressure (**Figure 1B**). Alpha distensibility has been shown to decrease with exercise induced PH, heart failure preserved ejection fraction and PAH resulting in a more dramatic effect on exercise induced increases in pressure.(11,35) In a subset of patients with RV pressure tracings with exercise, alpha distensibility significantly associates with exercise RV-PA coupling. (11) Alpha distensibility has been found to be modifiable in patients with heart failure reduced ejection fraction (HFrEF) (35) and exercise-induced pulmonary hypertension (17). When compared to placebo, 12 weeks of 12 weeks of sildenafil treatment significantly increased alpha distensibility by 24.6% in patients with HFrEF. (35) In HFrEF), the group with 12 weeks of sildenafil treatment had a 24.6% increase in alpha distensibility compared to those treated with the placebo.(35) In another mixed population of patients diagnosed with exercise induced pulmonary hypertension, Wallace et al. measured an increase in alpha distensibility from 0.69 ± 0.15 %/mmHg to 1.15 ± 0.27%/mmHg after 9.4 ± 4 months of vasodilator therapy. (17) The partial reversal of vascular distensibility with vasodilatory therapy suggests that alpha distensibility can be used as a marker of vascular beds response to therapy. In our study, patients with PAH did not have a significant change in alpha distensibility at their follow-up iCPET despite significant decrease in pulmonary vascular resistance and afterload (**Figure 5D**). At follow-up, participants on PH medications did have decreased pulmonary vascular resistance but it is possible that there was less of an effect on alpha distensibility due to the strength of therapy, degree of pulmonary vascular remodeling with the lower starting alpha distensibility of 0.13 [0.04-0.25] %/mmHg and decreased global PA compliance.

Alpha distensibility might be less modifiable in patients with advanced PAH and elevated PVR and therefore less effective at measuring therapeutic changes due to fairly advanced stages of pulmonary vascular remodeling. Alpha distensibility might act in a similar fashion as PA compliance in the resistance compliance curve (**Figure 5A and B**) (36) where compliance is less modifiable when PVR is high. In patients with exercise PH or mild-normal range of alpha distensibility, Langleben D. et al recently demonstrated there is still the ability for pulmonary capillary recruitment in addition for vascular distention during exercise.(37) The increase in alpha distensibility that Wallace et al. found in their cohort of exercise-induced PH cohort could potentially be explained by increased pulmonary capillary recruitment following pulmonary vascular therapy. In their cohort, resting mPAP decreased from 24.1 ± 1.7 mmHg pre-treatment to 21.6 ± 1.3 mmHg post treatment. While they found no change in resting PA compliance with treatment, they did show an increase in PA compliance at peak exercise. (17) We might speculate that in pulmonary capillary recruitment is significantly reduced in patients with advanced pulmonary vascular remodeling that is less modifiable by pulmonary vasodilator therapy. In our cohort of patients with advanced PAH, resting PVR decreased at follow-up (**Table 2**) but there were no significant changes in rest and exercise PA compliance and alpha distensibility. Based on the resistance-compliance relationship, there likely needs to be a large decrease in PA pressure and PVR for there to be a significant increase in alpha distensibility that is more difficult to achieve in patients with severe PAH with high PVR.

The Linehan distension model measures the distension of the resistive vessels and a key assumption in the model is that the resistive segments are fully recruited including the arteriolar, capillary and venular portions. Additional studies are needed to investigate the loss of capillary recruitment in advanced forms of PAH and whether it is modifiable with pulmonary vasodilatory therapy. Pulmonary vascular therapies that dilate the pulmonary vasculature but not reverse pulmonary vascular remodeling may not lead to significantly improved exercise induced changes in the vasculature (e.g recruitment). Thus, with vasodilatory therapy, vessels are more dilated at rest reducing resting resistance but if there is no change in vascular stiffness there will still be a significant exercise-induced increase in pressure based on alpha distensibility and the mPAP/Q slope. Alpha distensibility might be more modifiable in patients with advanced PAH with new PAH therapies like Sotatercept that more directly target pulmonary vascular remodeling through the BMP signaling pathway. (38)

#### Limitations

There are several limitations to the present study. The participants with a mean PA pressure less than 25 mmHg are not representative of a control population. Their alpha distensibility was 0.7 [0.39-0.76] %/mmHg suggest a degree of pulmonary vascular remodeling compared to a control population.(11). In fact, a few subjects had pressures between 20-25 mmHg and could be classified as having pulmonary hypertension under the new WSPH guidelines. (39) The cohort was retrospectively derived from the UA PH registry resulting in a cross-sectional view of participants with PAH at various stages of disease progression and therapeutic treatments. Age is another factor to consider with interpreting the exercise response and alpha distensiblity. Oliveira RK et al. demonstrated that subjects < 50 and ≥ 50 years old have different responses to exercise that result in different upper limits of normal.(33) In the present study, only eight participants were under the age of 50 years old and their PQ slope ranged from 1.4 to 21.6 at the index RHC. There was no significant correlation between age and alpha distensibility at the index RHC but there was one at the follow-up RHC (**Figure S3**).

Length of follow-up is another factor that could have had an influence with variable follow-up times from 3 months to 4.7 years. The change in alpha distensibility was not a function of the follow-up times but therapeutic strategies could have had a contributing factor in this retrospectively identified cohort.

## Conclusions

In this retrospective cohort analysis, we found limited changes in alpha distensibility with treatment in PAH patients despite a decrease in pulmonary vascular afterload (PVR and mPAP) at follow-up. The RShiny *iCPET calculator* standardizes and improves the reproducibility of the alpha distensibility calculations. Additional prospective studies are needed to investigate alpha distensibility as a clinical meaningful marker to study therapeutic effects in patients with advanced PAH and elevated PVR.

## Data Availability

The data is available upon request.

## Acknowledgements/Sources of Funding

This work was supported by the Archie Clifford and Clara Mabel Rentfrow Heart Disease Fund and the Mark and Emma Schiffman Research Fund from the University of Arizona Sarver Heart Center. This work was also supported by an AHA Career Development Award (19CDA34730039)

